# The effect of in-person primary and secondary school instruction on county-level SARS-CoV-2 spread in Indiana

**DOI:** 10.1101/2021.03.17.21250449

**Authors:** Gabriel T. Bosslet, Micah Pollak, Jeong Hoon Jang, Rebekah Roll, Mark Sperling, Babar Khan

**Affiliations:** Department of Medicine, Indiana University School of Medicine, Indianapolis, IN; School of Business and Economics, Indiana University Northwest, Gary, IN; Department of Biostatistics, Indiana University School of Medicine, Indianapolis, IN; Indiana University School of Medicine, Indianapolis, IN; School of Education, Indiana University Northwest, Gary, IN; Regenstrief Institute, Indianapolis, IN

**Keywords:** Public health, schools, education

## Abstract

**Background:** To determine the county-level effect of in-person primary and secondary school reopening on daily cases of SARS-CoV-2 in Indiana.

**Methods:** This is a county-level population-based study using a panel data regression analysis of the proportion of in-person learning to evaluate an association with community-wide daily new SARS-CoV-2 cases. The study period was July 12-October 6, 2020. We included 73 out of 92 (79.3%) Indiana counties in the analysis, accounting for 85.7% of school corporations and 90.6% of student enrollement statewide. The primary exposure was the proportion of students returning to in-person instruction. The primary outcome was the daily new SARS-CoV-2 cases per 100,000 residents at the county level.

**Results:** There is a statistically significant relationship between the proportion of students attending K-12 schools in-person and the county level daily cases of SARS-CoV-2 28 days later. For all ages, the coefficient of interest (β) is estimated at 3.36 (95% CI: 1.91—4.81; *p* < 0.001). This coefficient represents the effect of a change the proportion of students attending in-person on new daily cases 28 days later. For example, a 10 percentage point increase in K-12 students attending school in-person is associated with a daily increase in SARS-CoV-2 cases in the county equal to 0.336 cases/100,000 residents of all ages.

**Conclusion:** In-person primary and secondary school is associated with a statistically significant but proportionally small increase in the spread of SARS-CoV-2 cases.

## Introduction

Understanding the relationship between primary and secondary (K-12) school in-person instruction and community spread of severe acute respiratory syndrome coronavirus 2 (SARS-CoV-2) is limited, despite a wide range of approaches to learning since the beginning of the global outbreak. Some research suggests that the closure of schools helps to curb the spread of the virus.^1^ Other data have not demonstrated a measurable impact of school closure on COVID-19 spread. ^2,3^ Much of the literature about the reopening of schools and the spread of SARS-CoV-2 are case reports based upon contact-tracing studies.^4^ Many demonstrate adult to adult staff transmission to be a major contributor to spread.^5^ No studies to date have evaluated the role of K-12 in-person instruction to community spread of SARS-CoV-2. Seemingly this is the outcome of most interest given the transmission dynamics of the virus.

In the state of Indiana, decisions regarding when and how to reopen primary and secondary schools were made by local school corporations and health departments. Some school corporations elected for entirely in-person school, others started as virtual-only, and others had a hybrid system, which split children into cohorts and brought each cohort for in-person school on certain days of the week. In addition, most schools provided an elective virtual option. A statewide mask mandate enacted July 27, 2020 required students grades 3-12 and all teachers to wear a cloth face covering in situations where physical distancing was not possible when indoors in schools and on school buses.^6^ This decentralized approach to school reopening in the setting of statewide guidance regarding distancing, public gatherings, and masking presented a natural experiment for studying the effect of in-person school on SARS-CoV-2 spread.

This study evaluated the role of in-person instruction in primary and secondary schools on the community spread of SARS-CoV-2 in Indiana. Our primary hypothesis was that an increase in in-person primary and secondary school instruction would be associated with higher spread of SARS-CoV-2 cases in the community.

## Methods

This study used a panel data regression analysis to investigate the effect of the cumulative level of in-person instruction in schools on daily new SARS-CoV-2 cases at the county level in the state of Indiana. The study period of July 12, 2020 through October 6, 2020 was chosen to allow at least 6 weeks of data collection following school openings. Per the Indiana University-Purdue University Indianapolis Institutional Review Board, this study was not subject to human subjects research oversight due to the lack of individual-level data collection.

### Exposure Variable

The exposure variable is the proportion of students attending in-person instruction at K-12 public and charter schools in a county on a given day. This variable was constructed from the combination of school start dates and the estimated proportion of students attending in-person on the first day of school, which was assumed to remain constant during the study period. These data were obtained through a combination of public records and directly from individual school corporations.

In 2020 there were 399 public and charter school corporations in the 92 counties in Indiana.^7^ We limited our study to counties for which we were able to obtain estimates of the percentage of in-person instruction which covered at least 50% of enrolled students in those counties. As a result, our analysis includes 73 out of 92 (79.3%) counties in Indiana, accounting for 85.7% of the school corporations in the state, and 90.6% of the statewide student enrollment. One county, Monroe, was excluded as an outlier based on outlier analysis described in the **appendix**.

School corporations for which in-person instruction proportions could not be acquired were imputed using enrollment-weighted county averages to complete estimated in-person instruction proportions for all 343 school corporations in the 73 counties. In-person instruction was considered any method with students in-person one day per week or more. **Table 1** provides a summary of the characteristics of the counties and school corporations used in this study and within the state. 53% of the included school corporations were in counties in which greater than 50% of the students attended school in person, and 46% were in counties in which less than 50% attended in person.

**Table 1.** Summary of County and School Corporations in Indiana in relation to study outcomes and sample inclusion at the time of school reopening.

|  | State of<br>Indiana | Study<br>Counties | Study counties by proportion in-person<sup>†</sup> |  |  |  |
| --- | --- | --- | --- | --- | --- | --- |
|  |  |  | 0-25% | 25-50% | 50-75% | 75-100% |
| *<u>Counties</u>* |  |  |  |  |  |  |
| Number (%) | 92<br>(100%) | 73<br>(79.3%) | 2<br>(2.2%) | 8<br>(8.7%) | 5<br>(5.4%) | 58<br>(63.0%) |
| Population density (per sq-mile) | 178 | 201 | 345 | 752 | 324 | 118 |
| Median household income | \$53,915 | \$54,872 | \$52,584 | \$63,177 | \$52,936 | \$54,102 |
| Median age | 40.5 | 40.6 | 38.4 | 38.7 | 37.9 | 41.2 |
| *<u>Indiana Public and Charter School Corporations</u>* |  |  |  |  |  |  |
| Number (%) | 399<br>(100%) | 343<br>(85.7%) | 14<br>(4%) | 143<br>(36%) | 26<br>(7%) | 155<br>(39%) |
| Proportion of statewide enroll. | 100% | 92.0% | 4% | 38% | 13% | 35% |
| Avg. students per corporation | 2,634 | 2,816 | 3,262 | 2,824 | 5,079 | 2,390 |
| Proportion students in-person | - | 58.7% | 7.4% | 36.5% | 71.7% | 84.6% |
| Median instruction date start | 8/12 | 8/12 | 8/12 | 8/12 | 8/10 | 8/10 |
| *<u>SARS-CoV-2 case data</u>* |  |  |  |  |  |  |
| Total cases per 100,000 residents prior to 7/11 | 757 | 845 | 921 | 998 | 1001 | 629 |
| New daily cases per 100,000 residents between 7/12 to 10/6 | 13.1 | 13.3 | 21.4 | 12.0 | 14.3 | 13.3 |
| New daily cases per 100,000 residents between median start date and 10/6 | 8.6 | 9.1 | 24.0 | 12.0 | 15.3 | 14.3 |

\* Population density, median household income and median age reflect averages taken over included counties.
<sup>†</sup> Percentages do not add to 100% given that not all counties were included.

### Outcome Measures

Our primary outcome was the daily number of new SARS-CoV-2 cases per county per 100,000 residents based upon the date the test specimen associated with the positive result was collected. Daily SARS-CoV-2 case counts in each county were obtained via the Indiana State Department of Health data hub.^8^ For our secondary outcome, we limited these data to the 0-19 age group, which is based upon the date the positive result was entered into the Indiana State Department of Health system.^9^ This age range was selected because of reporting limitations to the Indiana Department of Health; we were unable to further stratify this group. County population data used for the per-capita outcome measures were obtained from the U.S. Census Bureau.^10^

### Statistical Analysis

Panel data regression analyses with fixed county and day effects were used to assess the relationship between the daily new SARS-CoV-2 cases per 100,000 residents and the proportion of students enrolled in-person 28 days earlier. The lag between the cumulative proportion of students utilizing in-person instruction and daily SARS-CoV-2 cases was set to 28 days (4 weeks); that is, we assumed 28 days between school start date and a potential effect on SARS-CoV-2 incidence. This period allows for time between any school-based contraction or spread of the virus and testing, as well as the potential for a child to infect others. This timing is consistent with other time-series studies of the effects of non-pharmacological interventions on SARS-CoV-2 incidence.^1,11^ Due to the significant day-of-the-week reporting differences, we smoothed the outcome measure using a 7-day average (current day and previous six days). The fixed county effects control for a wide range of time-invariant differences between counties, such as population density, socioeconomic and demographic factors, differences in mask and physical distancing restriction enforcement, ease of travel, and other factors. The fixed day effects capture time-varying state and national factors that commonly affect county SARS-CoV-2 infections such as statewide case rates, changes to state policies on social distancing and masks, and public awareness of SARS-CoV-2 severity. The model also controls for the 7-day average of daily county-level SARS-CoV-2 tests administered per 100,000 residents. Driscoll and Kraay’s robust standard errors^12^ were computed to account for cross-sectional dependence, autocorrelation and heteroskedasticity. Detailed description of the methods and justification for our modeling approach are described in the **appendix**.

We also conducted sensitivity analyses to examine the robustness of findings to both shorter (21-27 days) and longer (29-35 days) lag periods between school reopening and daily SARS-CoV-2 cases. We explored the sensitivity of our findings to two other methods of imputing missing school-level cumulative proportion of students in-person. In addition to the mean-value imputation employed for our primary analysis, we also imputed missing values at 0% in-person learning rates and 100% in-person learning rates with similar results. This analysis can be found in the **appendix**.

Analyses were performed using R (version 3.6.2, R Foundation for Statistical Computing, Vienna, Austria), and two-sided p-values of < 0.05 were considered statistically significant.

## Results

The statewide response to SARS-CoV-2 was coordinated by the governor and Indiana Department of Health. **Figure 1** outlines the study timeline in relation to daily new cases and statewide nonpharmacologic public health measures. The Institute for Health Metrics and Evaluation calculated Indiana’s mask adherence between 48% and 61% during the study period.^13^

**Figure 1.**
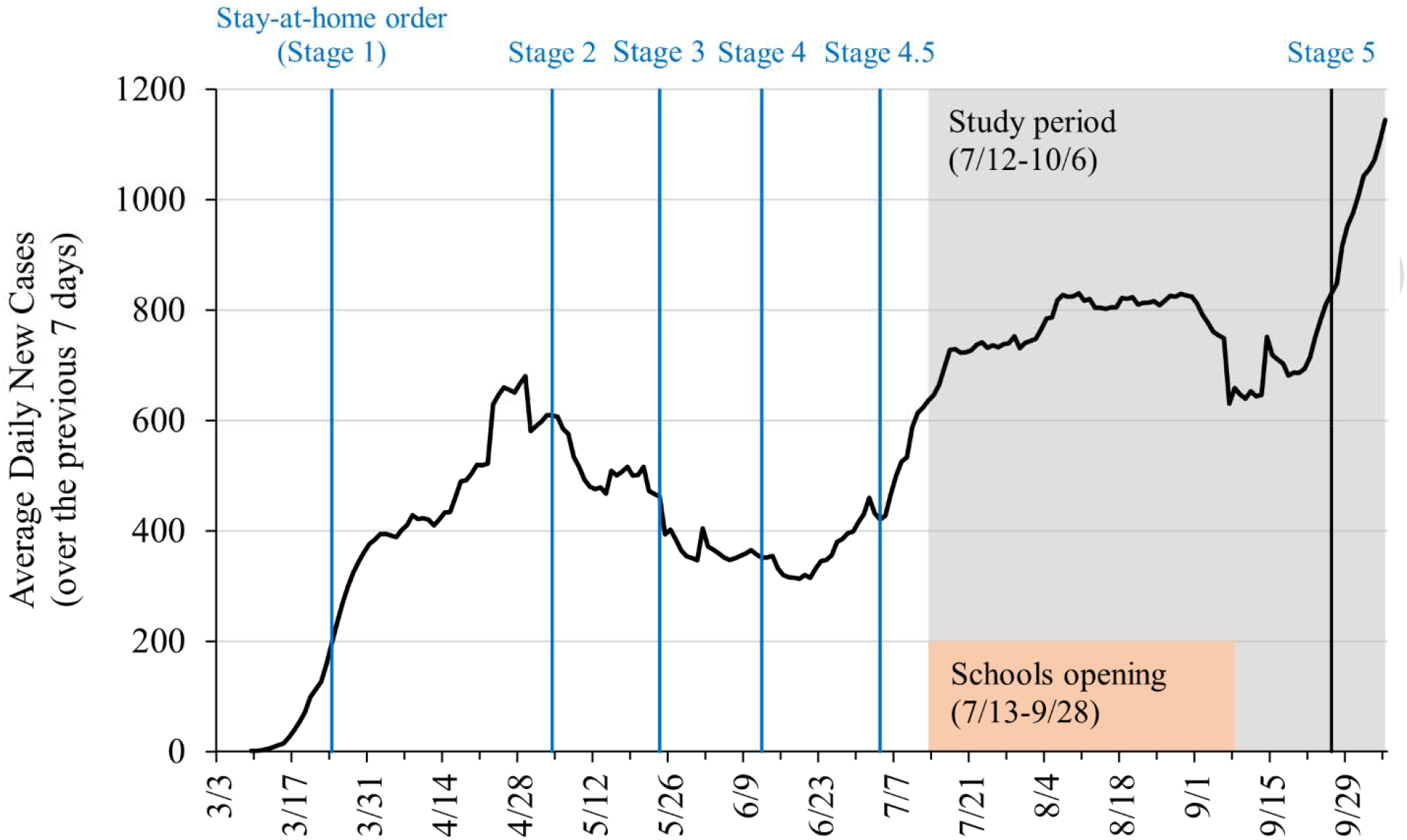
Average daily new SARS-CoV-2 cases over time for study counties including timeline of stages of Indiana re-opening, study period window and window of schools re-opening.

### Effect of in-person instruction on SARS-CoV-2 incidence

We found a positive and statistically significant relationship between the proportion of students attending K-12 schools in-person and the daily cases of SARS-CoV-2 per capita in that county 28 days later both for all ages and for ages 0-19 as shown in **Table 2**.

**Table 2.** Results of the panel data regression with county and day fixed effects using data from 73 counties in the state of Indiana. The main predictor is the county-level cumulative percentage of students utilizing in-person instruction.

| Outcome | $\beta$ (SE) | 95% CI | P-value |
| --- | --- | --- | --- |
| New daily SARS-CoV-2 cases per 100K | 3.36 (0.74) | [1.91, 4.81] | < 0.001 |
| New daily SARS-CoV-2 cases per 100K, ages 0-19 | 2.74 (0.56) | [1.63, 3.84] | < 0.001 |

\*Abbreviations: SE, standard error; CI, confidence interval; 100K, 100,000 residents.
\* $\beta$ represents the size of the effect a change in the proportion of students in-person has on new cases. Herein, new daily cases per capita = $\beta$ x (cumulative proportion of students in person) + other controls. For example, a 1% (0.01) increase in in-person instruction is associated with 0.036 additional daily cases per capita 28 days later (where 0.036 = 0.01 \* 3.36)

For all ages, the coefficient of interest (*β*) was estimated at 3.36 (95% CI: 1.91—4.81; *p* <0.001). This coefficient represents the effect of a change in the proportion of students attending in-person on new daily cases 28 days later. For example, a 10 percentage point increase in the proportion of K-12 students attending school in-person within a county (such as going from 50% in-person to 60% in-person or 0.5 to 0.6) was associated with a daily increase in SARS-CoV-2 cases in the county equal to (3.36 x 0.1) = 0.336 cases per 100,000 residents of all ages beginning 28 days later. To put this into terms of number of cases, during the period of this study the 73 counties included saw a total of 70,021 new SARS-CoV-2 cases among all ages, or an average of 805 cases per day. The estimated coefficient implies that a 10 percentage point increase in the proportion of students in-person would be associated with an additional 20 new cases per day (a 2.5% increase in average daily cases). The same decrease of in-person instruction would be associated with an equivalent decrease in new cases per day. However, because school corporations re-opened at different times during the period of the study these effects were introduced slowly and compounded over time. Using the estimated coefficient from our study, combined with the individual school district start dates and proportion of students in-person we can construct a profile of the number of SARS-CoV-2 cases that would likely have occurred under several different in-person instruction scenarios. For details on this construction, please see the **appendix**.

Applying the case change coefficient to the trajectory of new cases in Indiana over our study period allowed us to demonstrate the reduction or increase in new cases during this period under four potential scenarios (**Figure 2**). Had all school corporations in the included counties *decreased* their in-person instruction by 10 percentage points, there would have been an estimated 469 fewer total cases (−0.67% of cumulative new cases occurring during the study period) among all ages between July 12 and October 6. A reduction of in-person instruction by 25 percentage points would have resulted in an estimated 1,166 fewer total cases (−1.66%) among all ages during this period. Had all school corporations *increased* their in-person instruction by 10 percentage points during this time, there would have been an estimated 561 additional total cases (+0.80%) among all ages between July 12 and October 6. An increase of in-person instruction by 25 percentage points would have resulted in an estimated 1,147 additional total cases (+1.64%) among all ages during this period. Note that the results of the scenarios are not symmetric as not all school districts necessarily would have been able to increase or decrease their in-person instruction sufficiently in all scenarios due to their current in-person instruction levels.

**Figure 2.**
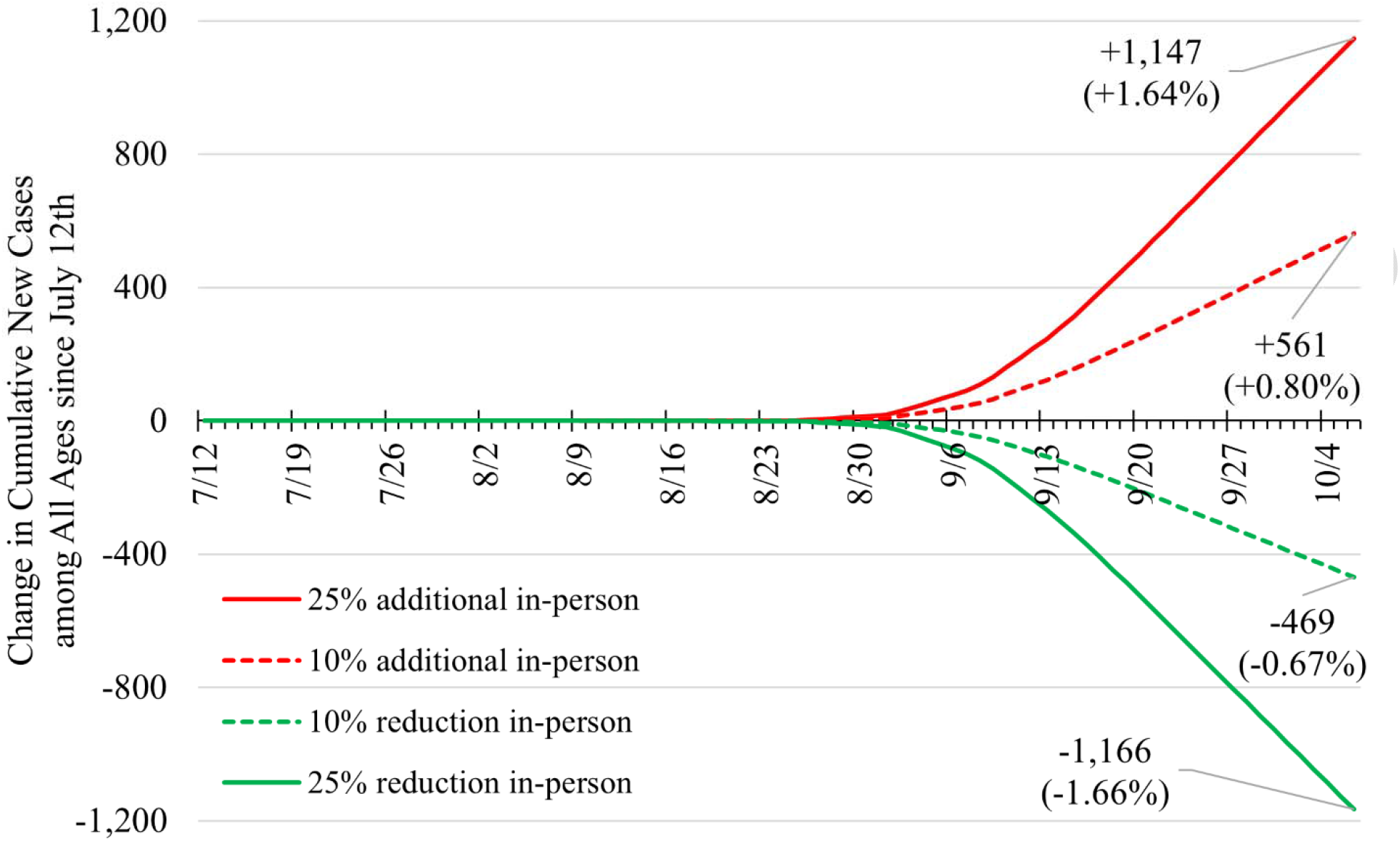
Predicted change in cumulative new cases between July 12^th^, 2020 and October 6^th^, 2020 among all ages under four scenarios of different levels of in-person instruction.

We found a similar relationship between in-person attendance and cases among those aged 0-19. For ages 0-19, the coefficient of interest was estimated at 2.74 (95% CI: 1.63—3.84; *p* < 0.001). During the study time there were 11,374 new SARS-CoV-2 cases among those aged 0-19. Based on the estimated coefficient, a 10-percentage point decrease in the proportion of K-12 students attending school in-person within a county was associated with a daily decrease in SARS-CoV-2 cases in the county equal to 0.274 cases (or a weekly decrease of 1.918 cases) per 100,000 residents aged 0-19 beginning 28 days later. **Figure 3** shows the reduction or increase in new cases among those aged 0-19 during the study period under the same four potential scenarios as in **Figure 2**.

**Figure 3.**
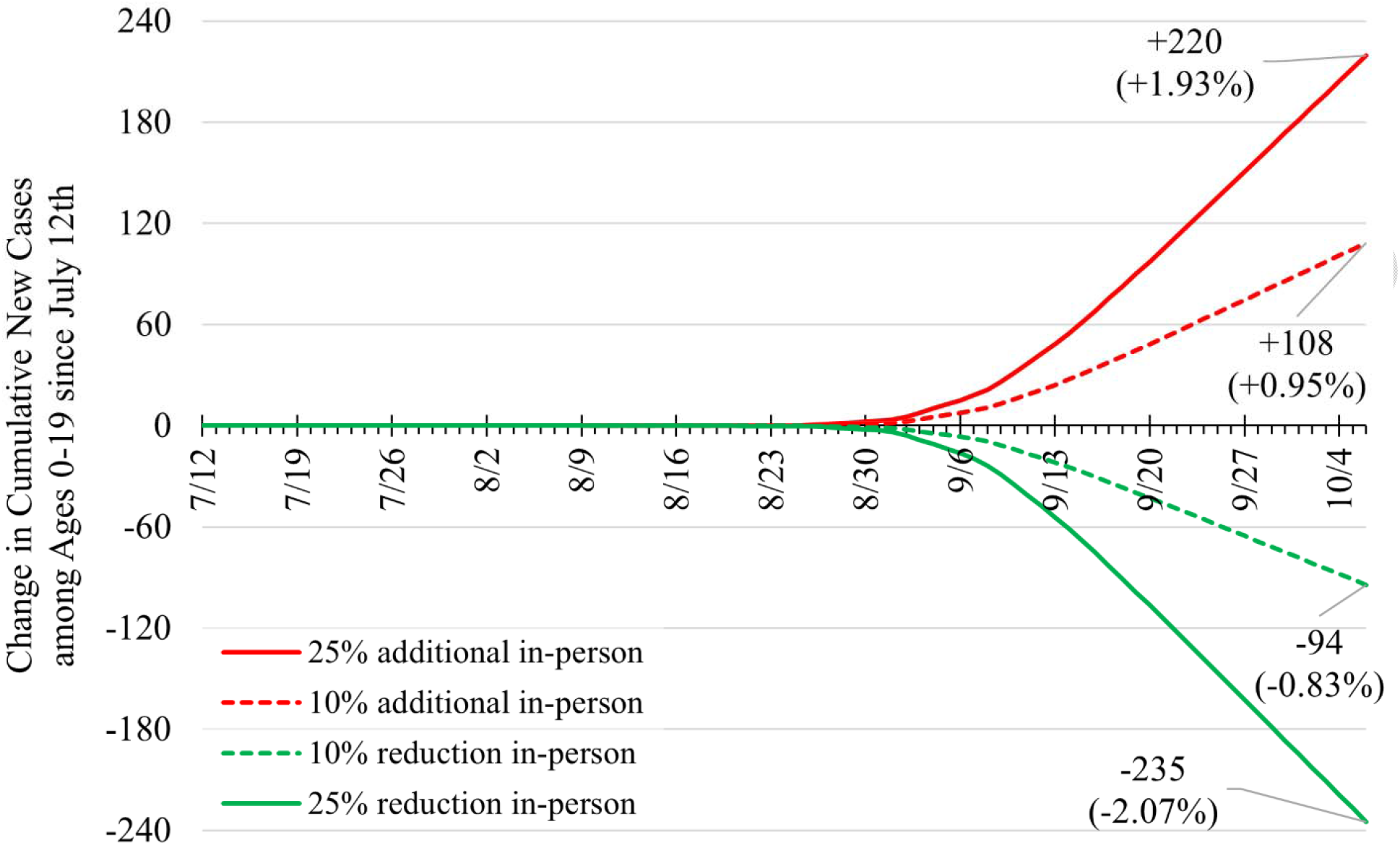
Predicted change in cumulative new cases between July 12^th^, 2020 and October 6^th^, 2020 among ages 0-19 under four scenarios of different levels of in-person instruction.

## Discussion

This study suggests that, in Indiana, primary and secondary school in-person instruction was associated with a statistically significant increase in the spread of SARS-CoV-2 cases both among those aged 0-19 and among the full population. However, this increase is relatively small in proportion to overall daily new cases. Specifically, this analysis suggests that 10% increase in the proportion of in-person primary and secondary instruction would increase total daily new cases by 0.80% of the total new daily cases 28 days later.

School reopening in Indiana occurred during a time of moderate community spread of SARS-CoV-2 (see **Figure 1**). Statewide, the number of daily new cases were 10 per 100,000 residents on July 12 and increased to 19 new cases per 100,000 residents slowly over the study period. The percentage of positive Polymerase Chain Reaction (PCR) tests was as high as 7.3% near the beginning of the study period and hovered to as low as 4.3%. This put Indiana into the “lower risk of transmission in schools” category for cases, and “moderate risk of transmission in schools” category for percentage of positive PCR tests, according to CDC guidelines, during the study period.^14^

There is reason to believe that opening schools with in-person instruction may not cause as much community spread as other indoor gatherings, although empiric data is mixed. Children seem to be less likely to have symptomatic infection with SARS-CoV-2^15^ and are less likely to be hospitalized due to COVID-19.^16^ Contact tracing studies suggest that, even among household contacts, children are less likely to spread SARS-CoV-2 to others.^5, 15^ Early modeling and data suggested that closing schools in Europe had little effect on spread of SARS-CoV-2,^4, 17^ and governmental reports from Europe suggest that schools played little role in community spread of SARS-CoV-2.^3^ A comparison between Sweden (schools remained open) and Finland (schools closed) suggested that schools did not have a measurable effect upon infections in school age children or in teachers.^2^ A recent case-control study of nearly 400 children in Mississippi comparing exposures of those with positive SARS-CoV-2 PCR tests to those with negative tests demonstrated that in-person school was not a risk factor for positivity.^18^ Additionally, a large evaluation of return-to-school efforts in Germany suggested that reopening of schools did not affect the transmission dynamics in that country.^19^

Paradoxically, other studies suggest that in-person schooling may lead to increased community transmission of SARS-CoV-2. One study suggested that the closure of schools in the United States during the initial surge of SARS-CoV-2 may have had a large and measurable impact on the transmission dynamics of SARS-CoV-2, and implied that closing schools may have saved tens of thousands of lives.^1^ In addition, a recent modeling study of country-wide interventions suggested that school closure had the effect of lowering the time-varying reproduction number and thereby reducing transmission of SARS-CoV-2.^20^

This study provides an explanation for why results from studies of in-person school instruction and spread of SARS-CoV-2 are mixed. It suggests that, while school is a large indoor gathering, which we know increases spread of SARS-CoV-2,^21^ the interactions of young people within a school building, when cloth face coverings and physical distancing practices are used, may not to drive community case transmission as much as might be expected.

## Limitations

This study has several limitations. First, we were not able to collect data on the proportion of students registered for in-person instruction for every school corporation in the counties included in the study. For the 73 counties included in the study, we were able to obtain proportion of in-person instruction data for school corporations covering at least 50% of the enrolled students in that county. For any school corporations in these counties for which we did not have data on the proportion of students registered for in-person instruction, we imputed this proportion to be equal to the enrollment-weighted county average of the corporations for which we had data. We believe this imputation is reasonable as the average difference between the proportion of students registered for in-person instruction at individual school corporations and the county average is only 4.6 percentage points, based on the counties where data make this comparison possible. Despite this, there may be school corporations for which we do not have data which have in-person proportions significantly different than the county average. To address this, the **appendix** provides regression results in which missing data are imputed first as 100% in-person and then 0% in-person, and the results are significant and similar in magnitude.

Second, these data do not account for any changes to in-person instruction that might have occurred following the first day of school. The study period (July 12, 2020 through October 6, 2020) was chosen to minimize potential widespread changes in instruction format, as most school corporations did not significantly alter their in-person policies during the first quarter. However, there may be a small number of students who changed between in-person learning and virtual learning during this period. Third, the linear nature of the model does not account for potential non-linear growth in cases that may result from spread due to in-person instruction. The effect of in-person instruction on daily new cases is assumed to be stationary when it is likely non-linear. However, given the relatively short study period, these non-linear effects should be minimal.

Lastly, we were unable to stratify school-age effects by grades or age. There is data to suggest that attack rates are directly proportional to the age of the child-essentially that younger children are less likely to become infected and possibly less likely to infect others.^22, 23^ This study was unable to assess any effect of age, other than for the broad 0-19 age group, on spread correlated with in-person school.

## Conclusion

This study suggests that primary and secondary school in-person instruction is associated with a statistically significant increase in the community spread of SARS-CoV-2 cases both among those aged 0-19 and the full population. However, this increase is relatively small in proportion to overall daily new cases.

## Data Availability

The data that support the findings of this study are available from the corresponding author, GB, upon reasonable request.

## Acknowledgements

The authors would like to acknowledge the Indiana State Department of Health, the Regenstrief Institute, and the Indiana Management Performance Hub for their transparent approach to data sharing that has helped lead the state of Indiana through the pandemic.

