## Supplement for "The effect of in-person primary and secondary school instruction on county-level SARS-CoV-2 spread in Indiana"

### Appendix

#### A. Empirical Model and Statistical Analysis

We employ a panel data regression model with fixed county and time effects to study the association between the cumulative level of in-person instruction in schools and daily new SARS-CoV-2 cases at the county level in the state of Indiana from July 12, 2020 through October 6, 2020 (an 87-day period). Specifically, we fit the following model:

$$Y_{it}= \beta_{1}X_{i,t-28}+\beta_{2}Z_{it}+\alpha_{i}+\gamma_{t}+u_{it}. (1)$$

$Y_{it}$ is the seven-day average of daily new SARS-CoV-2 cases per 100,000 residents in county i on day t ($t=1,.., 87$). The seven-day average (current date and previous dix days) is used to smooth out short-term fluctuations due to significant day-of-the-week reporting differences and highlight longer-term trends. In the secondary analysis, the outcome $Y_{it}$ is restricted to residents aged 0-19.

The primary independent variable $X_{i,t-28}$ denotes the daily cumulative proportion of students enrolled for in-person classes in county i on day $t-28$, where a 28-day lag between school start date and a potential effect on SARS-CoV-2 incidence is set to allow for time between any school-based contraction or spread of the virus and testing, as well as the potential for a child to infect others. This independent variable is calculated by aggregating school corporation-level data within each county as $X_{i,t-28}=\sum_{j=1}^{K_{i}} \theta_{ij}W_{ij,t-28}$, where $W_{ij,t-28}$ is the proportion of students attending in-person classes at $j$-th school corporation in county i on day $t-28$, $\theta_{ij}$ represents the enrollment size of the $j$-th school corporation as a proportion of the entire enrollment size in the $i$-th county, and $K_{i}$ denotes the number of school corporations in county i. Note that $W_{ij,t-28}$ equals 0 before the school start date (including the case of $t-28 \leq0)$, and equals the proportion of students attending in-person on the first day of school (school start date) and remain constant thereafter. The estimated in-person instruction proportions were acquired for 220 out of the 343 school corporations, accounting for 79.3% of all enrollment in these counties. School corporations for which in-person instruction proportions could not be acquired were imputed using enrollment-weighted county averages. To minimize bias, the model includes data from 73 counties, for which we were able to obtain estimates of the proportion of in-person instruction which covered at least 50% of enrolled students in those counties. While this was the lower cut-off, for many counties we had substantially more data. For 28 of these 73 counties we obtained estimates of the proportion of in-person instruction for every school corporation. For 44 of these 73 counties we obtained estimates for 75% or more of all school corporations by total enrollment.

In addition, the model (1) controls for $Z_{it}$, the seven-day average of daily county-level SARS-CoV-2 tests administered per 100,000 residents. The county fixed-effects $\alpha_{i}$ control for a wide range of time-invariant differences between counties, such as population density, baseline hospital capacities, median income, insurance coverage rates and other socioeconomic and demographic factors. The fixed day effects $\gamma_{t}$ control for time-varying state and national factors that commonly affect county SARS-CoV-2 infections such as statewide case rates, changes to state policies on social distancing and masks and public awareness of SARS-CoV-2 severity. $u_{it}$ are idiosyncratic error terms.

For estimation, we first re-write model (1) as:

$Y_{it}= {\beta_{0}+\beta}_{1}X_{i,t-28}+\beta_{2}Z_{it}+\rho_{2}C_{2}+ \cdots+\rho_{73}C_{73}+\delta_{2}T_{2}+ \cdots+\delta_{87}C_{87}+u_{it}, (2)$

where $C_{i}$ (i$=2,\ldots, 73$) and $T_{t}$ (t$=2,\ldots, 87$) are binary county and day dummy variables, respectively. Then, the regression coefficients of the re-written model (2) are estimated using the using the plm function in the R plm package.^1^ Driscoll and Kraay's robust standard errors^2^ were computed to account for cross-sectional dependence, autocorrelation and heteroskedasticity.

#### B. Justification of the Proposed Modeling Approach

In this section, we provide additional justification for the use of panel data regression model with county and time fixed effects to study the association between the cumulative level of in-person instruction in schools and daily new SARS-CoV-2 cases at the county level. Significant results of the F test for two-ways effects (*p* < 0.001) and Hausman test^3^ (*p* < 0.001) indicate that the fixed effects model is preferred to the ordinary least squares model and random effects model, respectively. In other words, we employed the fixed effects model to correct for potential omitted variable bias caused by: i) unobserved time-invariant heterogeneities across the entities ii) unobserved county-invariant heterogeneities across time; iii) and correlation between the observed explanatory variables and the unobserved effect. A series of Lagrange Multiplier tests^4^ for county and time fixed effects (separately and combined) further justifies the validity of our modeling approach (all p-values < 0.001). Significant results of Paseran CD test^5^ (*p* = 0.010), Breusch-Godfrey test^6^ (*p* < 0.001) and Breusch-Pagan test^7^ (*p* < 0.001) respectively suggest the existence of cross-sectional dependence, autocorrelation and heteroskedasticity in the fixed effects model; this justifies the use of Driscoll and Kraay's robust standard errors for valid statistical inference. Similar test results and justification were derived for the use of fixed effects model with the outcome restricted to residents aged 0-19 (secondary analysis).

Alternative modeling approaches include interrupted time series (IPT) and segmented regression analyses. Although these approaches are useful for evaluating the longitudinal effects of interventions through regression modelling, their use is limited in our setting where “intervention” is introduced and accumulated gradually/continuously over time as schools successively start in-person instruction.

#### C. Outlier Analysis

We performed outlier analysis to detect individual counties that might disproportionally influence the overall estimate of the fixed effect panel regression model (1). Specifically, the Cook’s distance was calculated for each data point by leaving out the point and fitting the regression on the remaining data points. Figure S1 presents the histogram of Cook’s distances averaged by county under model (1) where the outcome is county-level daily new SARS-CoV-2 case per 100,000 residents. Monroe county clearly stands out as the most influential county with the largest average Cook’s distance (0.0019) above the threshold (0.0010). Pike county is the second influential county (average Cook’s distance = 0.0015). Figure S2 presents the average Cook’s distance by county under model (1), this time restricting the outcome to residents aged 0-19. We see that Monroe county is the single most influential county, with average Cook’s distance much higher than those of other counties as well as the threshold. These findings are sensible given that Monroe county, with largest college campus in the state, had a large testing mitigation strategy and had an outlier number of tests and positive cases found in the student population during our study. Monroe county also has a relatively high student to non-student population ratio compared to other counties with major college campuses. We thus excluded Monroe county from the analysis.

#### D. Sensitivity Analysis

In the first set of sensitivity analyses, we examined the robustness of findings to both shorter (21-27 days) and longer (29-35 days) lag periods between school reopening and daily SARS-CoV-2 cases. The results are presented in Table S1. We see that the effects are relatively small and non-significant when the lag is set to 21 days (3 week), suggesting that this is not a long enough window period for the effect of in-person learning on the daily new SARS-CoV-2 cases at the county level to manifest among all residents ($\beta=1.29;SE=0.68;P=0.058$). However, we are able to find relatively strong and consistent effects of in-person learning across longer lags ranging from 25 to 35 days. Specifically, the strongest and highly significant effects are found when the lag is set to either 28 (among all residents: $\beta=3.36;SE=0.74;P<0.001$) or 31 days (among all residents: $\beta=3.32;SE=0.61;P<0.001$). Similar findings are reported when the outcome is restricted to residents aged 0-19.

In the second set of sensitivity analyses, we explored the sensitivity of our findings to two other methods of imputing missing school-level cumulative proportion of students in-person: 1) imputing missing values with 0% in-person learning rates (assuming 100% e-learning rates); and 2) imputing missing values with 100% in-person learning rates. Results are presented in Table S2. When the missing values were imputed with 0% in-person learning rates, we found very strong effects of in-person learning on the daily new SARS-CoV-2 cases both among all residents ($\beta=4.81;SE=0.94;P=0.028$) and among those aged 0-19 ($\beta=5.27;SE=1.36;P<0.001$). When the missing values were imputed with 0% in-person learning rates, we found results similar to those of our primary analysis presented in the main manuscript (where missing values were imputed by enrollment-weighted county averages).

#### E. Constructing Scenarios in Figure 1 and Figure 2

To construct the four scenarios in Figure 1 and Figure 2, we use the following procedure. First, at the school corporation level, we adjust the estimated proportion of students attending by the appropriate percentage point change according to the scenario (+25%, +10%, -10%, -25%). Not all corporations are able to adjust fully. For example, if a school corporation has 80% of students attending in-person then they can increase by 10 percentage points (to 90%) but can only increase 20 out of the full 25 percentage points, as the maximum attending in-person is 100%. Using the adjusted proportion of students attending in-person, we then aggregate school corporations to the county level to construct the cumulative proportion of students attending in-person on a given day in that county, taking care to account for different start dates by school corporations. Combining this with the appropriate estimated coefficient ($\beta$**)**, we can then determine the effect on daily new SARS-CoV-2 case per 100,000 residents in the county. For example, if a county has a single school corporation that increases its in-person attendance by 10 percentage points then beginning 28 days after the school start date, daily new SARS-CoV-2 case per 100,000 residents in the county will increase by 0.1 x 3.36 = 0.336. We then aggregate the daily effects across counties and populations to generate the total effect on daily new SARS-CoV-2 case per 100,000 residents in all counties in the sample, which is presented in Figure 1 for all ages and in Figure 2 for ages 0-19 only.

**Figure S1.** Histogram of average Cook’s distance by county under model (1), where the outcome is county-level daily new SARS-CoV-2 case per 100,000 residents. The red vertical line at 0.001 denotes the threshold above which the data point is considered an outlier.

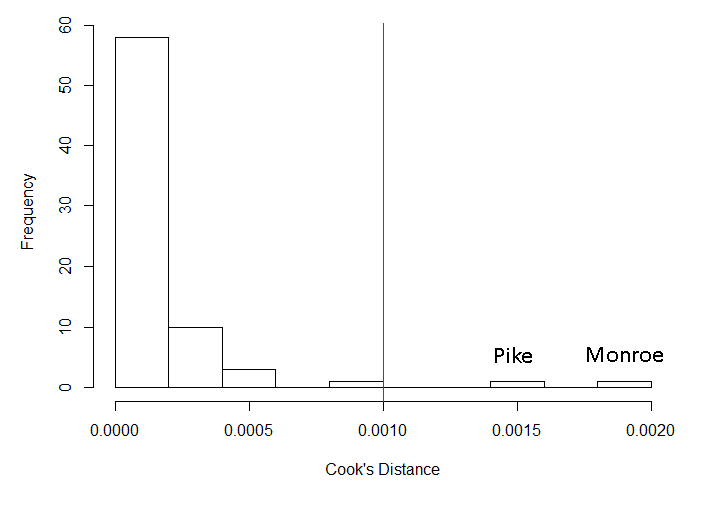

**Figure S2.** Histogram of average Cook’s distance by county under model (1), where the outcome is county-level daily new SARS-CoV-2 case per 100,000 residents aged 0-19. The red vertical line at 0.001 denotes the threshold above which the data point is considered an outlier.

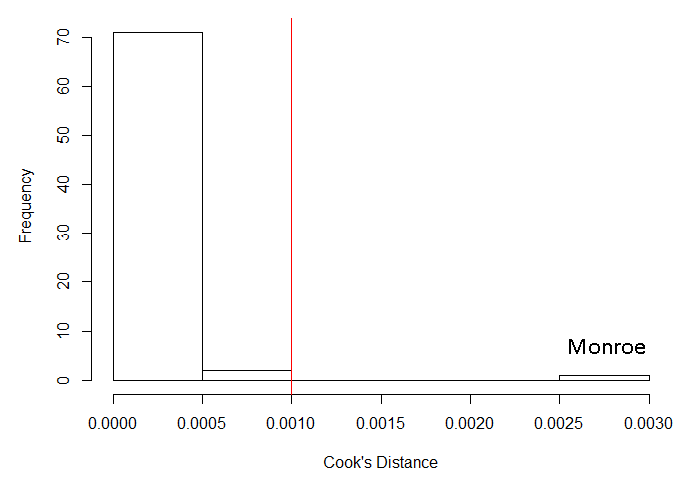

**Table S1.** Results of the panel data regression with county and day fixed effects using data from 73 counties in the state of Indiana for different lags of the main predictor, which is the county-level cumulative proportion of students utilizing in-person instruction.

| Lag | Outcome | $\boldsymbol{\beta}$ (SE) | 95% CI | P-value |
| --- | --- | --- | --- | --- |
| 21 days | New daily SARS-CoV-2 cases per 100K | 1.29 (0.68) | [-0.04, 2.63] | 0.058 |
|  | New daily SARS-CoV-2 cases per 100K, ages 0-19 | 1.78 (0.52) | [0.76, 2.79] | 0.001 |
| 25 days | New daily SARS-CoV-2 cases per 100K | 2.65 (0.80) | [1.07, 4.23] | 0.001 |
|  | New daily SARS-CoV-2 cases per 100K, ages 0-19 | 2.02 (0.60) | [0.85, 3.19] | 0.001 |
| 28 days | New daily SARS-CoV-2 cases per 100K | 3.36 (0.74) | [1.91, 4.81] | < 0.001 |
|  | New daily SARS-CoV-2 cases per 100K, ages 0-19 | 2.74 (0.56) | [1.63, 3.84] | < 0.001 |
| 31 days | New daily SARS-CoV-2 cases per 100K | 3.32 (0.61) | [2.12, 4.52] | < 0.001 |
|  | New daily SARS-CoV-2 cases per 100K, ages 0-19 | 2.94 (0.46) | [2.03, 3.85] | < 0.001 |
| 35 days | New daily SARS-CoV-2 cases per 100K | 2.84 (0.65) | [1.57, 4.11] | < 0.001 |
|  | New daily SARS-CoV-2 cases per 100K, ages 0-19 | 2.12 (0.61) | [0.92, 3.32] | 0.001 |

- Abbreviations: SE, standard error; CI, confidence interval; 100K, 100,000 residents.

- $\boldsymbol{\beta}$ represents a change in new daily SARS-COV-2 cases per 100K residents given 1 unit (100 percentage point) increase in the county-level cumulative proportion of students utilizing in-person instruction, holding other control variables fixed.

**Table S2.** Results of the panel data regression with county and day fixed effects using data from 73 counties in the state of Indiana under two methods for imputing missing school-level cumulative proportion of students in-person: 1) imputing missing values with 0% in-person learning rates; and 2) imputing missing values with 100% in-person learning rates. The main predictor is the county-level cumulative percentage proportion of students utilizing in-person instruction.

| Impute^a^ | Outcome | $\boldsymbol{\beta}$ (SE) | 95% CI | P-value |
| --- | --- | --- | --- | --- |
| 0% | New daily SARS-CoV-2 cases per 100K | 4.81 (0.94) | [2.96, 6.65] | < 0.001 |
|  | New daily SARS-CoV-2 cases per 100K, ages 0-19 | 5.27 (1.36) | [2.61, 7.93] | < 0.001 |
| 100% | New daily SARS-CoV-2 cases per 100K | 3.32 (0.94) | [1.48, 5.16] | < 0.001 |
|  | New daily SARS-CoV-2 cases per 100K, ages 0-19 | 2.51 (0.86) | [0.83, 4.19] | 0.003 |

- Abbreviations: SE, standard error; CI, confidence interval; 100K, 100,000 residents.

- $\beta$ represents a change in new daily SARS-COV-2 cases per 100K residents given 1 unit (100 percentage point) increase in the county-level cumulative proportion of students utilizing in-person instruction, holding other control variables fixed.

^a^Two methods for imputing missing school-level cumulative proportion of students in-person: 1) imputing missing values with 0% in-person learning rates (0%); and 2) imputing missing values with 100% in-person learning rates (100%).
